# Fusing a Bayesian case velocity model with random forest for predicting COVID-19 in the U.S.

**DOI:** 10.1101/2020.05.15.20102608

**Authors:** Gregory L. Watson, Di Xiong, Lu Zhang, Joseph A. Zoller, John Shamshoian, Phillip Sundin, Teresa Bufford, Anne W. Rimoin, Marc A. Suchard, Christina M. Ramirez

**Affiliations:** Department of Biostatistics, Fielding School of Public Health, University of California, Los Angeles, California, United States of America; Department of Epidemiology, Fielding School of Public Health, University of California, Los Angeles, California, United States of America; Departments of Biomathematics and Human Genetics, David Geffen School of Medicine, University of California, Los Angeles, California, United States of America

## Abstract

Predictions of COVID-19 case growth and mortality are critical to the decisions of political leaders, businesses, and individuals grappling with the pandemic. This predictive task is challenging due to the novelty of the virus, limited data, and dynamic political and societal responses. We embed a Bayesian nonlinear mixed model and a random forest algorithm within an epidemiological compartmental model for empirically grounded COVID-19 predictions. The Bayesian case model fits a location-specific curve to the velocity (first derivative) of the transformed cumulative case count, borrowing strength across geographic locations and incorporating prior information to obtain a posterior distribution for case trajectory. The compartmental model uses this distribution and predicts deaths using a random forest algorithm trained on COVID-19 data and population-level characteristics, yielding daily projections and interval estimates for infections and deaths in U.S. states. We evaluate forecasting accuracy on a two-week holdout set, finding that the model predicts COVID-19 cases and deaths well, with a mean absolute scaled error of 0.40 for cases and 0.32 for deaths throughout the two-week evaluation period. The substantial variation in predicted trajectories and associated uncertainty between states is illustrated by comparing three unique locations: New York, Ohio, and Mississippi. The sophistication and accuracy of this COVID-19 model offer reliable predictions and uncertainty estimates for the current trajectory of the pandemic in the U.S. and provide a platform for future predictions as shifting political and societal responses alter its course.

**Author summary:** COVID-19 models can be roughly classified as mathematical models that simulate disease within a population, including epidemiological compartmental models, or statistical curve-fitting models that fit a function to observed data and extrapolate forward into the future. Bridging this divide, we combine the strengths of curve-fitting statistical models and the structure of epidemiological models, by embedding a Bayesian nonlinear mixed model for case velocity and a machine learning algorithm (random forest) into the framework of a compartmental model. Fusing these models together exploits the particular strengths of each to glean as much information as possible from the currently available data. We also identify the velocity of log cumulative cases as an excellent target for modeling and extrapolating COVID-19 case trajectories. We empirically evaluate the predictive performance of the model and provide predicted trajectories with credible intervals for cumulative confirmed case count, active confirmed infections and COVID-19 deaths for each of the 50 U.S. states. Combining sophisticated data analytic methods with proven epidemiological models offers an empirically grounded strategy for making realistic predictions and quantifying their uncertainty. These predictions indicate substantial variation in the COVID-19 trajectories of U.S. states.

## Introduction

Rapid spread of SARS-CoV-2 virus across the planet has precipitated a global pandemic, infecting millions and killing hundreds of thousands. Governments around the world have undertaken unprecedented interventions aimed at curtailing the spread and lethality of the virus. These interventions and more recently proposals for relaxing them have relied heavily on predictions of COVID-19 case growth and mortality.

COVID-19 prediction models can be roughly classified as mathematical models that simulate disease within a population or statistical models that fit a function to observed data and extrapolate forward into the future. We will discuss the features of both types of models. Most COVID-19 models are compartmental models [1–61], a type of mathematical model used by epidemiologists to simulate infectious disease epidemics for over a century. Compartmental models divide a population into mutually exclusive compartments that denote disease status and supply a set of differential equations that define the flow of the population between compartments [62]. Traditionally they are named after their compartments with the SIR (susceptible-infectious-recovered) [63] and SEIR (susceptible-exposed-infectious-recovered) models classic examples.

In an infectious disease compartmental model, *S*(*t*) is the number of susceptible individuals at time *t*, and new infections are represented by the flow of individuals out of the S compartment. This is governed by the first derivative of *S*(*t*) with respect to time, *dS*(*t*)*/dt*. Classic SIR and SEIR models express this as proportional to the product of *S*(*t*), *I*(*t*), and a rate constant *β*,

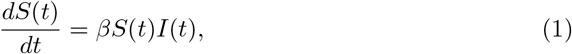

where *I*(*t*) is the number of infectious individuals at time *t*. The rate *β* is often interpreted as disease transmissibility and may be expressed as a function of the reproductive number *R*_0_—the expected number of individuals infected by an infectious person—and contact rates between individuals. It may also be normalized in Eq 1 by division with the total population size.

The simplest approach for simulating infections is to assume a value for *β* or its constituent parts from the literature or other prior information [1–17]. While this is convenient, the predictive accuracy can suffer. Another approach that has been used by other studies is to estimate *β* (or a related quantity) by fitting a statistical model or other optimization procedure to observed data [18–39]. This empirical approach can make these models more realistic, but they still may be limited in their ability to accurately model the COVID-19 pandemic. Disease transmission rates in COVID-19 have changed substantially over time depending upon the political and societal responses and possibly other factors [54]. As a result, modelers operating within this framework often resort to modeling transmission rate changes by applying an adjustment factor that modifies transmission rates upward or downward in a somewhat ad hoc manner.

This has motivated modeling efforts that allow the disease transmission rate to vary over time, i.e., replacing *β* in Eq 1 with *β*(*t*) [40–50]. This is a promising approach, but to be useful for forecasting, estimates of *β*(*t*) must extrapolate beyond the observed data to describe transmission at unobserved times and not simply interpolate the observed data, which is straightforward with a flexible model. Two studies have paired machine learning algorithms with their COVID-19 compartmental models to model time-varying effects, which is a promising approach at least when inference on the inputs to *β* is not required. Yang et al. fit a long short-term memory neural network to data from the 2003 SARS outbreak adjusted by the output of their SEIR model [45]. Dandekar and Barbastathis augmented their compartmental models with a neural network that models time-varying transmission by estimating intervention efficiency from reported data as a function of time [42].

Recovery, death, and other states (e.g., hospitalization) may be incorporated into the model as separate compartments. Solutions to the differential system provide values for each compartment at each time, allowing for easy joint modelling of disease states once their derivative is specified. This is an advantage of compartmental models over many other approaches, which may require separate models for each quantity.

A number of agent-based COVID-19 models have been developed or adapted from influenza pandemic models to simulate the individuals of a population and their interactions [64–68]. This provides a mechanism for modelling interventions that target contacts between individuals and does not assume the population exists in homogeneous compartments as compartmental models generally do, but also requires a number of assumptions to be made on the behavior and interactions within a population as well as the infectivity of COVID-19.

Serial growth models for COVID-19 simulate an epidemic by expressing the number of new infections at a given time as a weighted sum of new infections on previous days usually scaled by the reproductive number, which may be time-varying [69–73]. The weights are sampled from a probability distribution defining the amount of time between an individual being infected and infecting another person. Deaths or other outcomes may be modelled as a second step, for example using a negative binomial model that predicts daily deaths conditional on the number of infections in recent days [70].

Statistical models often eschew deterministic population dynamics and fit the observed data as a function of time and possibly other covariates in a regression (or equivalent) framework. Log-linear [74], generalized Richards [75], ARIMA [76,77], exponential [78], Gaussian CDF [79], and logistic [80–82] models, which all accommodate the generally sigmoidal shape of the cumulative infection count that is often observed in epidemics, as well as various other models [83–86] including machine learning algorithms [87–89] have been proposed for COVID-19. Murray et al. and Woody et al. take similar approaches for modeling COVID-19 deaths using the error function (ERF) [90,91]. Modeling deaths is appealing, because COVID-19 deaths have been more reliably reported than infections. However, because deaths lag infections by some amount of time, it may not enable projections to incorporate the latest information on disease spread.

Within the framework of a statistical (or other regression-like) model, it is easier to fit observed data, assuming an appropriate functional form is selected, but it may be challenging to accurately project the future trajectory of an epidemic. Time-varying covariates like mobile phone tracking data [91], Google trends [89,92], and social media [93] are easily incorporated into such a model and may be quite predictive, but they are often unknown in the future, requiring assumptions to be made regarding their future values when forecasting. Such approaches can only model one outcome (e.g., infections or deaths) and additional steps must be taken to predict other quantities. Here we present an approach to projecting COVID-19 cases and deaths that employs sophisticated data analytic methods to learn transition functions for a compartmental model. We fit a Bayesian mixed model to the velocity of COVID-19 case growth, providing location-specific trajectories that extrapolate well within a full probability model that includes uncertainty quantification. We use a random forest algorithm for the death transition function that learns the relationship between COVID-19 cases and population characteristics to predict deaths. The remaining sections of this paper lay out the SIRD compartmental model, the Bayesian mixed model for case velocity, the random forest death model, and close with results and a discussion.

## Materials and methods

### The SIRD Compartmental Model

We model the spread and progression of COVID-19 through a population using a SIRD compartmental model named after the four compartments into which it partitions the population: *S* for susceptible, *I* for infectious, *R* for recovered, and *D* for dead. The number of population members in each compartment is a function of time, *t*, and these functions are linked by a system of ordinary differential equations (ODEs) that govern the flow of the population through the different disease states:

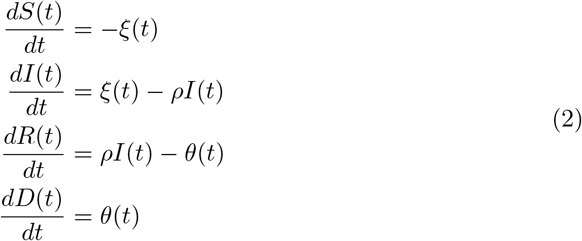

Figure 1 graphically depicts the SIRD model with arrows between compartments indicating possible transitions between compartments. Only deaths due to COVID-19 are permitted within this framework under the assumption that ignoring other causes of death, as well as the influx of new susceptible persons through birth or immigration, will not substantially alter inference in the short term.

**Figure 1.**
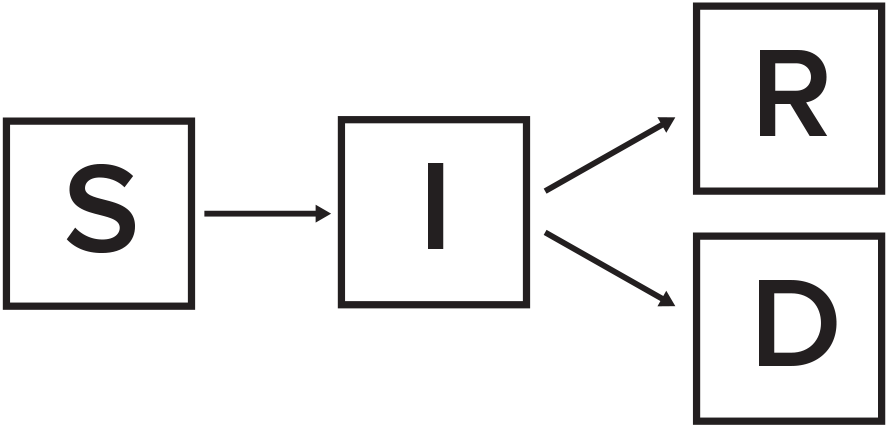
**The SIRD Model.** Each of the four compartments quantifies the number of population members with that disease status: S for susceptible, I for infected, R for recovered and D for dead. The arrows indicate possible transitions between disease states.

The transition rates between compartments are determined by the functional forms and parameter values in Eq 2. Given these and initial conditions for the system, *S*(*t*_0_), *I*(*t*_0_), *R*(*t*_0_), and *D*(*t*_0_), the system of ODEs in Eq 2 is deterministic, but in general does not accommodate analytical solutions. Consequently, we compute numerical solutions using the lsoda solver in the deSolve package [94] of R [95].

Due to the novelty of the SARS-CoV-2 virus and a desire to empirically ground the compartmental model, we fit transition functions that can vary in time and incorporate covariates and other information. The transition between S and I is determined by *ξ*(*t*), which describes the number of individuals becoming confirmed COVID-19 cases and is presented in the next section. In many locations there is no reliable data on the transition of individuals out of I into R, except for hospitalized patients in some places. We follow the standard SIR approach and model transition out of I with a rate parameter *ρ* that is the inverse of the number of days a person is expected to be infected. The destination of individuals transitioning out of I is determined by the death model *θ*(*t*).

Initial conditions for the model were constructed by stepping the system through the observed case count data and then projecting forward. This approach should be more accurate than simply beginning the simulation on the last observed day, because the distribution of cases into compartments I, R, and D is not observed.

### Case Velocity Model

COVID-19 case counts across U.S. states and culturally similar European nations have exhibited relatively consistent trajectories. Once community transmission had been established, cases grew exponentially until social distancing and lock down interventions were enacted, which have gradually curbed case growth. We investigate the dynamics of COVID-19 case growth by modeling the velocity, i.e., the first derivative with respect to time, of the log cumulative case count. This is the instantaneous rate of new cases to cumulative cases at a given time, and is very similar to the reproductive number. Calculating the reproductive number at a particular time, however, requires knowing the number of active infections. There is currently no reliable data on this, as most infections resolve on their own outside of a clinical or otherwise supervised setting in which their transition from active case to recovered might be recorded. The velocity of log cumulative cases on the other hand is readily estimated from the data and presents itself as an appealing target of analysis.

A very crude estimate of the derivative can be obtained using first differences, but smoothing allows for more precise estimates, as calculating the derivative requires some notion of function smoothness [96]. We estimate the velocity by fitting a cubic spline to the observed log cumulative case count and then evaluating its derivative at the observed time points. Since there is relatively little noise in the cumulative accounts, we assume any uncertainty introduced by this procedure is negligible.

Figure 2 depicts the velocity for U.S. states with the horizontal axis enumerating time since 100 or more confirmed cases were reported in that location, a milestone that proxies for the establishment of community transmission. Community transmission or its proxy is a sensible time point for data alignment, because there is substantial variation observed in the length of time between the detection of the first cases in a location and the acceleration of cases accompanying community transmission. This variation likely reflects both the possibility of containing a small number of initial cases and the increased uncertainty accompanying small samples.

**Figure 2.**
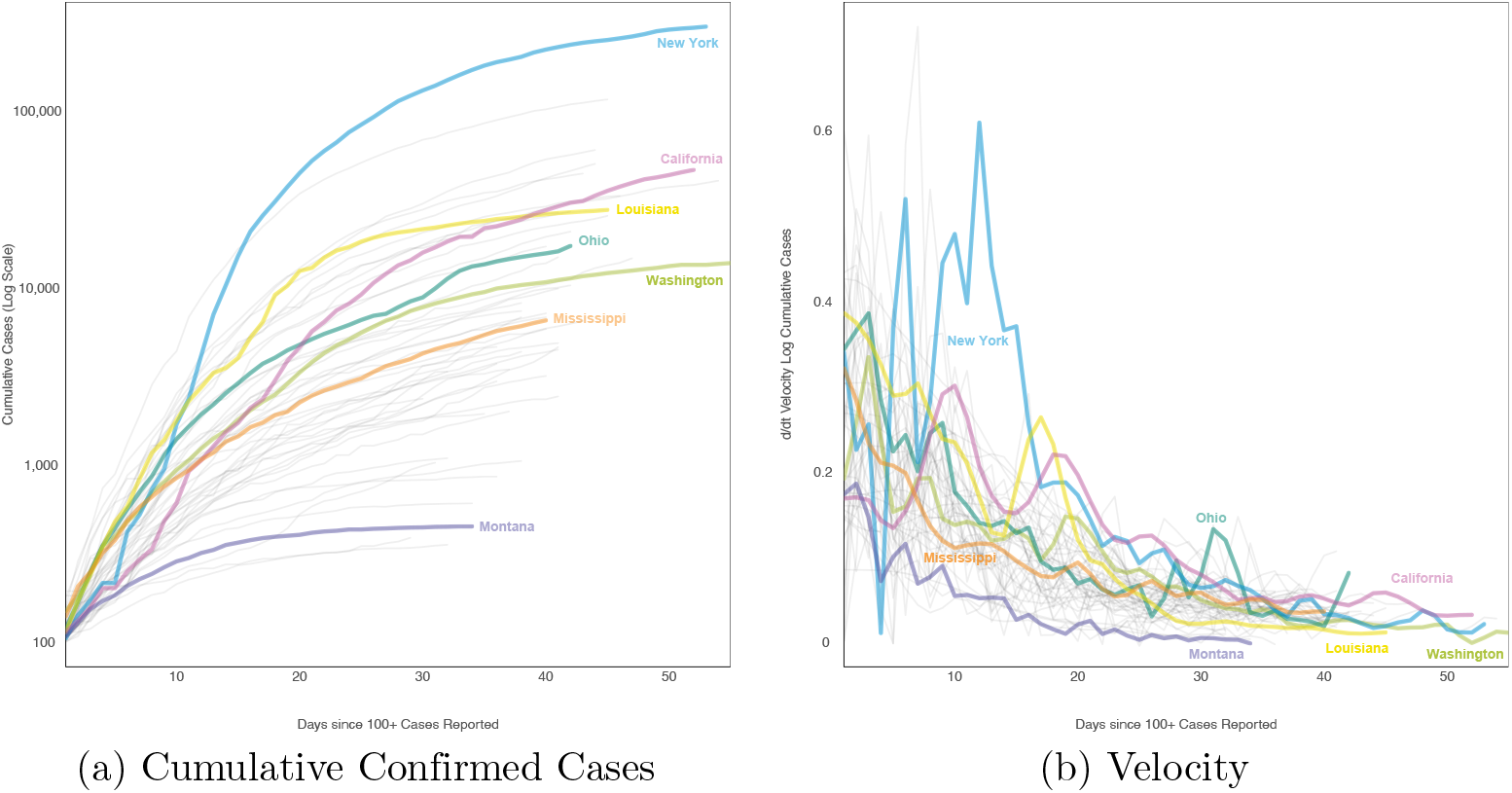
**Log Cumulative Cases and Its Velocity.** The log cumulative confirmed case count (a) and its velocity (b), i.e., first derivative with respect to time, for each of the 50 U.S. states since 100 or more confirmed cases were reported.

The velocity of a cumulative function cannot be negative, since cumulative functions are monotonically increasing. Consequently, we employed a log link to map velocity to the entire real line and modeled its mean with a linear predictor. We selected a Bayesian mixed model to obtain location-specific estimates of the trajectory. Location-specific random effects for the intercept and slope borrow strength across locations for more precise estimates while accommodating individual variation. Borrowing strength can be particularly helpful for estimating the trajectory of locations with smaller populations or less advanced outbreaks.

Let *u_i_*(*t*) denote the cumulative case count for location *i* at time *t*, and *y_i_*(*t*) the first derivative with respect to time of its log transformation, i.e., *y_i_*(*t*) = *d* log *u_i_*(*t*)*/dt*. Log-transformed velocity is modeled as a linear combination of random effects and Gaussian noise,

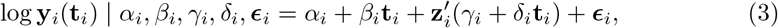

where y*_i_*(t*_i_*) is a vector of length *n_i_* denoting the velocity evaluated at times t_*i*_ = (*t_i_*_1_*,…, t_ini_*)^*0*^; z_*i*_ = (*x_i_*_1_*,…, x_ini_*)^*0*^is a vector indicating whether each time occurred after intervention, i.e., *z_ij_* = *I*(*t_ij_ ≥ π_i_*); *α_i_* is the random intercept, *β_i_* the random slope, *γ_i_* the random effect for intervention, and *δ_i_* is the random slope for intervention. The elements in the error vector_*i*_ are assumed to be independently Gaussian distributed with mean zero and precision (inverse variance) *τ_ij_*, where *j* = 1*,…, n_i_*. In vector notation,

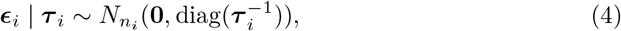

where *N_ni_* is an *n_i_* dimensional Gaussian distribution, its mean 0 is a vector of zeros, and its covariance matrix is diagonal with elements 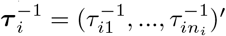.

Obvious heteroskedasticity is apparent in the observed case counts with variation decreasing with time. To account for this and to allow for the variance to differ between locations, the precision vector for location *i, τ_i_*, was modeled as linear in time with location-specific random effects,

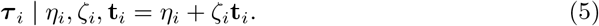

Each of the velocity and precision random effects were assigned Gaussian priors,

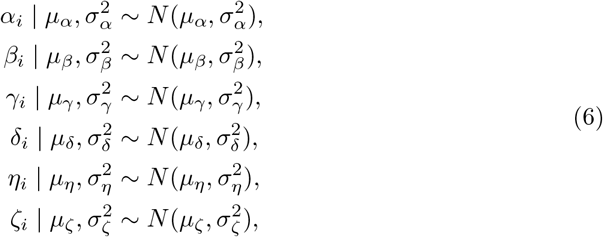

with means *µ_α_, µ_β_, µ_γ_, µ_δ_, µ_η_, µ_ζ_*, and variances 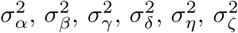. These means were themselves given Gaussian hyperpriors. The prior mean and variance values used for the predictions presented here are listed in S2 Table of the supplemental material.

Posterior inference was conducted via Markov chain Monte Carlo (MCMC) simulation using JAGS 4.3.0 and the `R2jags` [97] package of R. Three chains of 500,000 iterations each were run after a burn in of 10,000 iterations. Visual inspection of parameter trace and autocorrelation plots indicated the chains mixed well.

The posterior estimate of the location-specific lognormal fit at the last observed time point was converted into a transition function for use in the compartmental model. Let *a_i_* + *b_i_t* denote the linear predictor for location *i* for the last observed time point. For locations in which an intervention was enacted (most locations), *a_i_* and *b_i_* are the post-intervention intercept and slope, while the pre-intervention line was used for non-intervening locations,

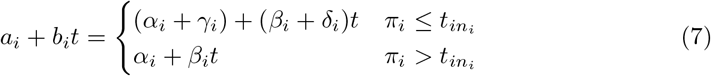

where *t_ini_* is the final observed time point for location *i*. The lognormal model for *d* log *u_i_*(*t*)*/dt* implies that

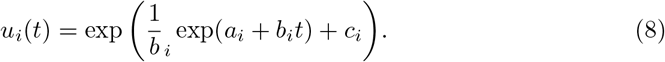

The MCMC procedure described above provided samples from the posterior distributions of *a_i_* and *b_i_*, but does not uniquely identify *c_i_*, because the value of a function cannot be deduced from its derivative alone. We empirically estimate *c*^(*m*)^_*i*_ by minimizing a squared loss function defined over the observed cumulative count at location *i* for each posterior sample, *m* = 1*,…, M*,

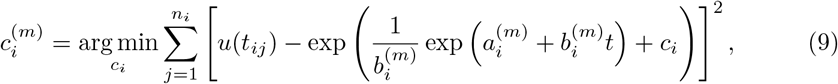

where the addition of a superscript^(*m*)^to a parameter denotes its *m*-th posterior sample. This procedure provides a posterior distribution for *c_i_*, and by extension for *u_i_*(*t*). Noting that 1 *− S*(*t*) gives the cumulative number of cases at time *t* in the compartmental model described above, we set *dS_i_*(*t*)*/dt* = *− ξ_i_*(*t*) = *−du_i_*(*t*)*/dt*. The posterior mean or median of *−du_i_*(*t*)*/dt* could be used as an estimate of *ξ*(*t*), but simply plugging in this single function into the SIRD model would ignore the uncertainty of this estimate. To incorporate this uncertainty explicitly into the SIRD model, we run the model separately for each posterior sample, giving a distribution of rate transition functions, *ξ_i_*(*t*)^(1)^*,…, ξ_i_*(*t*)^(*m*)^. Accounting for uncertainty is particularly important for our application, because COVID-19 predictions without interval estimates quantifying uncertainty may lead decision makers to place undue confidence in their accuracy.

### Death Model

Infected individuals either recover or die, which corresponds to a transition from compartment I to R or D. The transition rate out of I is the inverse of the expected number of days a person is infected. Various estimates for this have appeared in the literature. Attempting to synthesize these various accounts while incorporating some of the uncertainty, we sample *ρ^−^*^1^ for each run of the compartment model from a Gaussian distribution with mean 14 and standard deviation one.

The number of individuals transitioning from I to compartment D on a particular day is predicted using a random forest model trained on the numbers of cases and deaths reported by the individual U.S. states. Random forest is a heuristic machine learning prediction algorithm that combines a large number of regression or classification trees into an ensemble [98]. It is a very commonly used algorithm known to perform well at a variety of predictive tasks [99].

Let *d_ij_* denote the number of dead reported in location *i* on day *j*, where days are indexed for each location from the first day on which 100 or more cumulative confirmed cases were reported in that location. Let w_*ij*_ = (*w_ij_*_1_*,…, w_ijp_*)^*0*^denoting the vector of *p* covariates for location *i* on day *j*. The conditional expectation of *d_ij_* given covariates w_*ij*_ is modeled as a random forest, i.e., as an ensemble of bootstrapped regression trees,

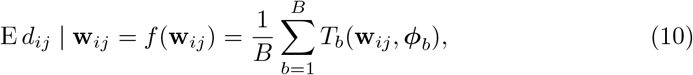

where *b* = 1*,…, B* indexes bootstrap samples of the training data, and *T_b_*(w_*ij*_, *φ_b_*) is a regression tree trained on the *b*-th bootstrap sample that relates covariate vector w_*ij*_ to parameters *φ_b_*. The model was fit using the `randomForest` package [100] of R using the default parameter values for the number of trees (500) and the number of covariates considered for each recursive split of the covariate space (floor(*p/*3)).

Figure 3 lists the covariates included in the model and their importance scores. Age, sex and comorbidity have been consistently reported in the literature as important risk factors for COVID-19 mortality. Even in the U.S. where testing has been limited, we expected that COVID-19 deaths on a particular day would be highly related to the number of cases reported on preceding days. Consequently the number of newly reported COVID-19 cases in location *i* on days *t −* 1*,…, t −* 14 were included as 236 covariates for predicting deaths on day *t*.

**Figure 3.**
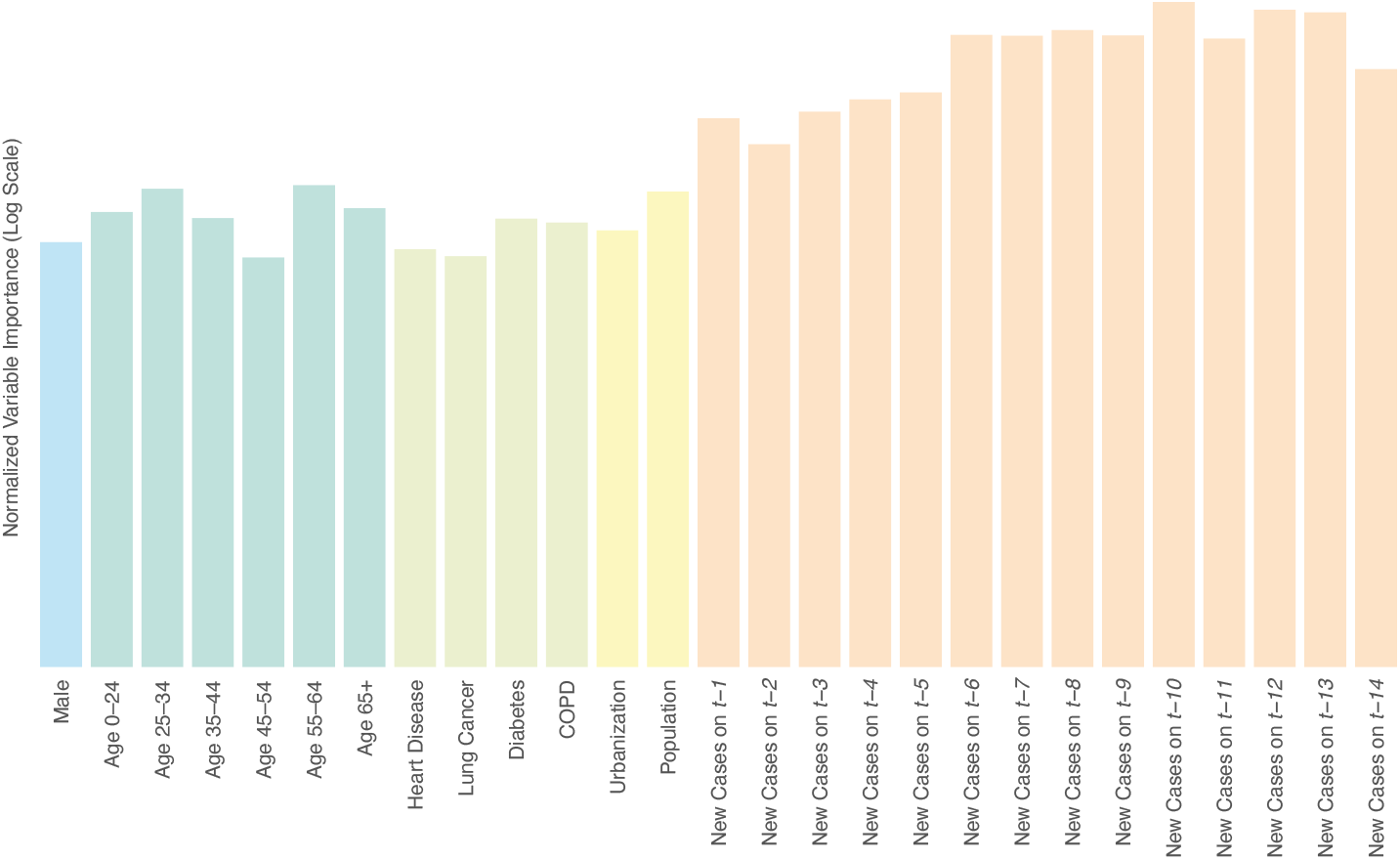
**Death Model Covariate Importance.** Covariate importance scores on the log scale for the random forest death model as the mean decrease in MSE associated with permutation of the variable’s values.

Covariate importance scores were computed using permutation variable importance. Briefly, permutation importance can be thought of as the decrease in predictive accuracy (in terms of mean squared error (MSE)) between the original model and when each variable is randomly permuted thus obscuring any signal it has with the outcome variable. The idea is that if a covariate is important in terms of prediction, obscuring its signal should result in a decrease in predictive accuracy. The most important of the lagged cases was that at *t −* 10, indicating that the model was able to discern a lag time between positive tests and COVID-19 fatalities. Additional lagged cases beyond 14 days were not included in the model, even though they may have been informative, because each additional lagged day reduces the number of available training observations at each location.

Fitting the model to data collected through April 29, 2020, resulted in a very high out-of-bag *R*^2^of 0.90. This is an overly optimistic estimate of prediction error, due to the within-location and temporal dependence of the data [101], but more significantly due to the lagged case counts being very informative covariates. Lagged cases were far more important than the demographic characteristics, which is not surprising considering the very strong relationship between testing positive for COVID-19 and dying of COVID-19, especially in the early days of the pandemic in the U.S., when testing was quite limited. The random forest predictions were capped at a percentage of the new cases to avoid unrealistically high death predictions, which can occur when there are relatively few new cases. This upper bound was set to be equivalent to a 15% case fatality rate for the first 30 days of the epidemic and reduced to 7% subsequently, with the higher initial death rate motivated by the relative severity of early confirmed cases due to limited testing.

### Predictive Accuracy

We assessed the predictive accuracy of our model by training it on case and death counts collected through April 15, 2020, and predicting through April 29. We quantified prediction error for each state on each day using the mean absolute scaled error (MASE) of the posterior median number of new cases and deaths. MASE is computed by dividing the mean absolute prediction error by the in-sample mean absolute error of a naive random walk forecast,

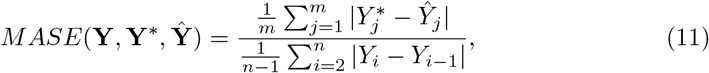

where Y = (*Y*_1_*,…, Y_n_*)^*0*^is the training data outcome, 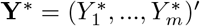 is the observed outcome in the evaluation set and 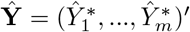 is the prediction for Y^*∗*^to be evaluated [102]. A MASE of one indicates that the predictions were on average equally accurate to the mean accuracy of a random walk forecast in the training data. A useful feature of MASE for our purposes is its scale invariance, which makes comparisons of predictive accuracy between states with epidemics on different scales more meaningful. Figure 4 depicts the distribution of MASE across states for cases and deaths over the two-week prediction period. The overall MASE for cases was 0.4 and for deaths was 0.32. As expected, the mean and variance of the MASE increased for both cases and deaths across the prediction period, as the time between the end of the training data and the date of the forecast increased. On April 29, a full two-weeks beyond the training data, the MASE for cases and deaths was still well below one, an encouraging sign for the reliability of our predictions. A longer evaluation period would allow the estimation of forecasting error farther into the future, but the brief period for which COVID-19 data are available makes this currently infeasible.

**Figure 4.**
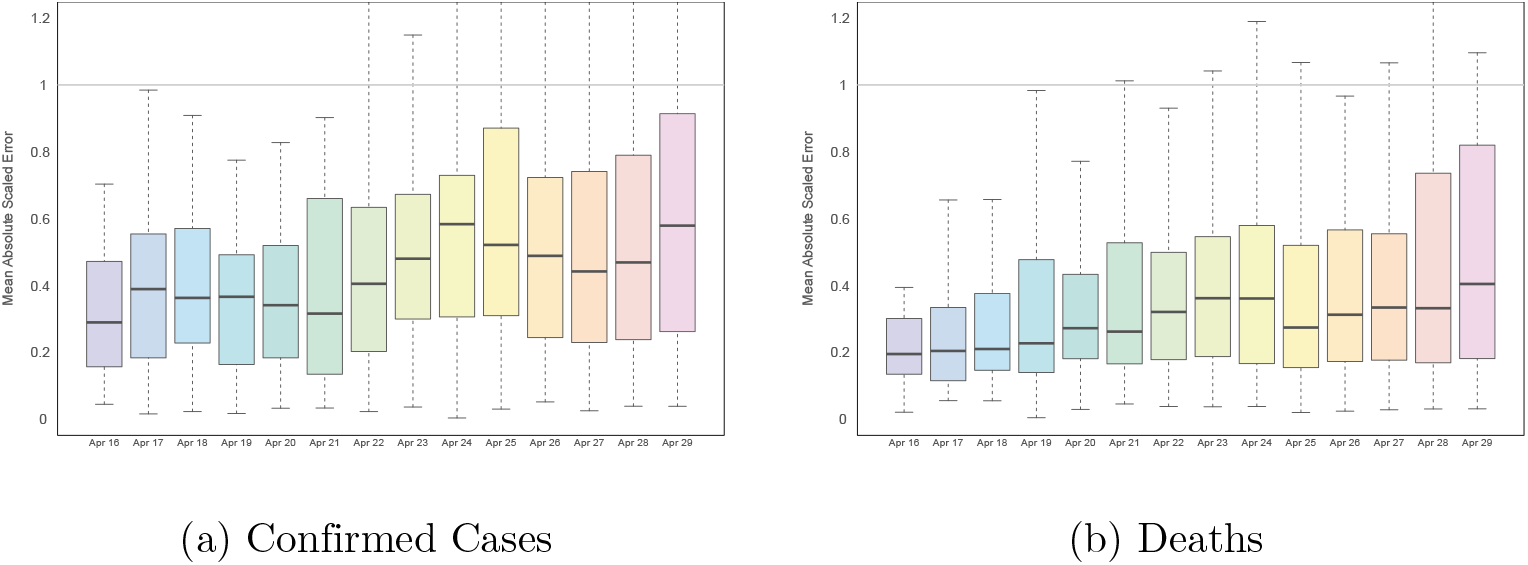
**Predictive Accuracy.** The distribution of MASE across all 50 states on each day of the 14-day prediction period for new confirmed cases and deaths.

## Results & Discussion

Infections and deaths were projected through July 1, 2020, for all 50 states. Figure 5 depicts median predicted cumulative confirmed cases as well as active confirmed infections and daily death counts for New York, Ohio, and Mississippi. These three states were selected as examples, because they are diverse in their population size, geography, political alignment, demographics, and in the progression of their COVID-19 epidemics. The equivalent figures for the remaining 47 states are included in the supplemental material S1 Fig.

**Figure 5.**
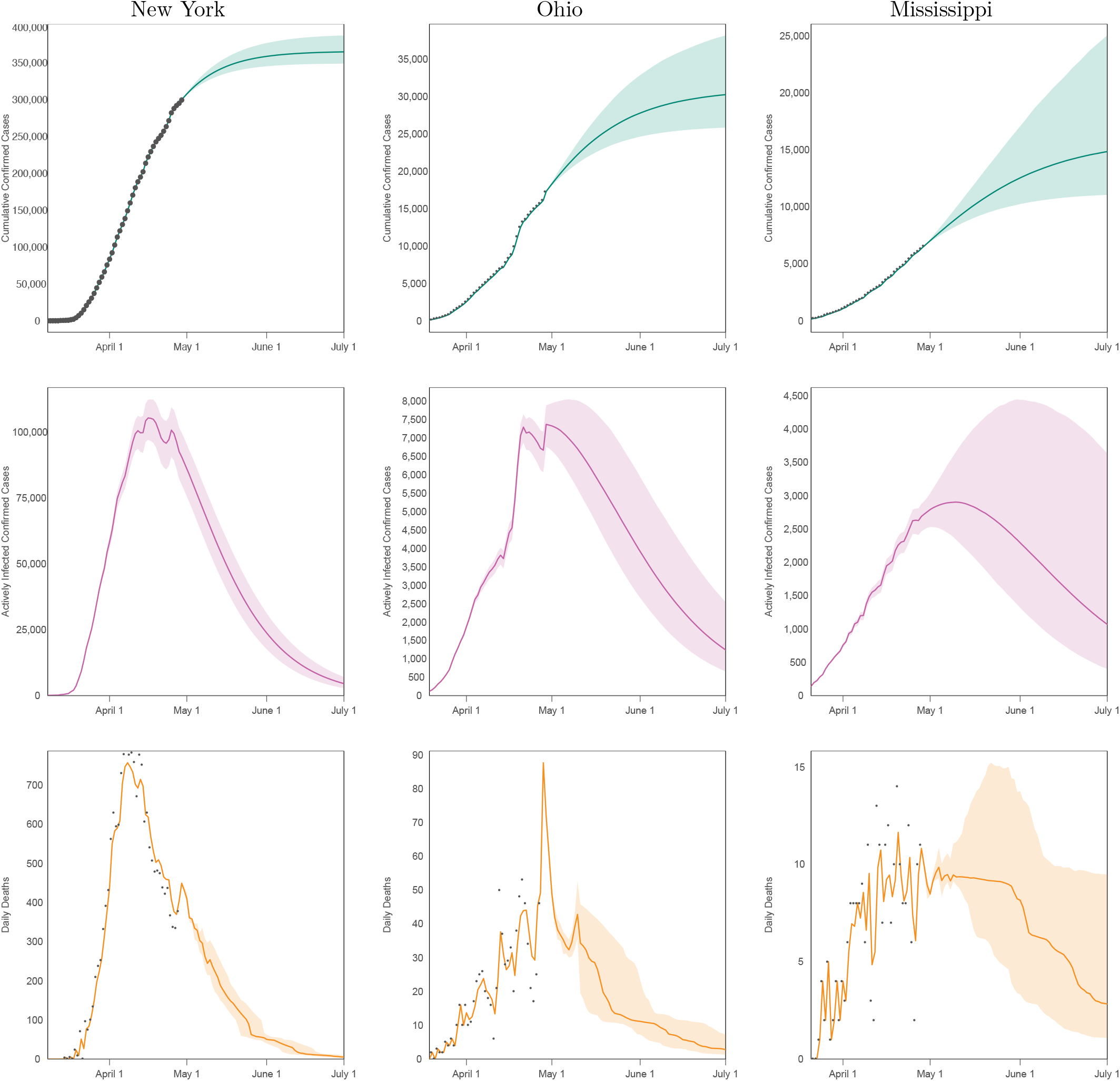
**Predicted Cumulative Cases, Active Infections & Deaths.** Projected cumulative case count, active confirmed infections, and daily deaths through July 1, 2020, for New York, Ohio, and Mississippi. The grey dots indicate observed data, which are not available for active infections.

These trajectories depend upon the ongoing societal and political response to the pandemic. In particular the current trajectory downward in case growth is due in no small part to the substantial interventions that have been undertaken across the U.S. The incorporation of covariates into the case growth and mortality models allow for alternate trajectories to be explored, which is the subject of ongoing research. The cumulative case counts eventually plateau for each state as its case velocity decreases toward zero. The predicted daily death counts are not smooth because the random forest algorithm averages many discontinuous segments into a prediction.

New York, especially New York City with its large, dense population, has been the epicenter of the largest COVID-19 outbreak in the United States with over 300,000 confirmed cases by late April. Initial exponential case growth was slowly curbed by public interventions, leading to a consistent decrease in case velocity and peaks in active cases and deaths in mid April. Case growth being well past its peak translates into a plateauing cumulative case curve and a relatively narrow interval estimate compared to states that peaked more recently or have yet to peak.

Like many other states, Ohio has had many fewer cases than New York with approximately 18,000 cases and appears to be peaking near the end of April. Its interval estimates are relatively wider than New York, because there is less uncertainty in the estimated trajectory farther past the peak. Ohio also exhibits more relative variation in its daily death counts than New York because of the smaller number.

Mississippi with fewer than 7,000 cases through the end of April illustrates the estimated trajectories of a relatively rural, Southern state that has not yet peaked. With cases farther from their plateau, there is correspondingly more relative uncertainty in its trajectory and a much wider range of dates over which its peak may occur.

The figures include 95% credible intervals around the median indicating that 95% of simulation results fell within this region. These intervals are not true credible intervals in the Bayesian sense, because random forest is not a probability model. Nevertheless, they represent a reasonable account of model uncertainty, as they incorporate credible intervals from the Bayesian case model and uncertainty around the duration of illness.

Despite the many strengths of the current approach, it is not without limitations. The projections produced here assume states continue upon their current trajectories. Changes in policy interventions, for example, could result in substantial deviation from this. Projecting outcomes under different or changing intervention scenarios is the subject of ongoing work.

Considering COVID-19 cases and death over large areas can obscure variation on a smaller scale. It is possible for a generally positive trajectory at the state-level to mask a burgeoning outbreak in some locale within the state until that outbreak contributes sufficiently many cases to influence the state-wide trajectory. A more granular approach that models COVID-19 at a finer resolution may be able to identify such an outbreak earlier.

There is substantial interest in estimating the proportion of the population that has or will have recovered from COVID-19 in the hopes that these individuals have acquired at least temporary immunity to the virus and can be the vanguard to economic recovery. Since we focus on modeling confirmed cases and deaths, our model does not predict the true number of recovered individuals. It is well known that, especially in the U.S., confirmed cases are a substantial undercount for the true number of COVID-19 infections. As a result, estimating the number of recovered individuals requires additional information beyond predictions of confirmed cases and deaths. Attempts to quantify recovery using serology testing are underway in the U.S. and elsewhere.

There is residual temporal autocorrelation in the case velocity not captured by the random effects for intercept and slope in the mixed model. We expect this has minimal impact on our posterior trajectories, as we are primarily concerned with the trend in mean over time, but incorporating a more sophisticated approach for temporal dependence could be used to explicitly model this autocorrelation.

Finally, one could consider more elegant methods for incorporating lagged case counts into a death model than simply inserting them as covariates into random forest. However, many approaches to lag estimation are only good retrospectively and thus are insufficient for the current task.

## Data Availability

The data underlying the results presented in the study are available from https://github.com/COVID19Tracking/covid-tracking-data.

## Supporting Information

**S1 Fig. State Predictions.** Projected cumulative case count, active confirmed infections, and daily deaths through July 1, 2020, for each of the 50 U.S. states. 352

**S2 Table. Parameter Values and Prior Distributions.**

## Acknowledgments

We thank Donatello Telesca, Jay J. Xu, and Ian Frankenburg (University of California, Los Angeles) for their helpful comments and assistance, and Private Health Management whose support helped make this work possible.

